# Where do Brazilian dental students seek information about COVID-19?

**DOI:** 10.1101/2020.08.24.20179614

**Authors:** Maria Gerusa Brito Aragao, Francisco Isaac Fernandes Gomes, Camila Siqueira Silva Coelho, Letícia Pinho Maia Paixão de Melo, Silmara Aparecida Milori Corona

## Abstract

**Objective:** We investigated where Brazilian dental students seek information about COVID-19 by a self-administered web-based questionnaire.

**Methods:** A social network campaign on Instagram was raised to approach the target population. The dental students responded to a multiple-response question asking where they get information about COVID-19. The possible answers were government official websites or health and education institutions websites, TV Programs, professors, social media, scientific articles, other health professionals, and family members. The data were analyzed by descriptive statistics and frequency distributions of responses were evaluated by gender, age, type of institution, and year of enrollment.

**Results:** A total of 833 valid responses were received. The main source of information used by the dental students were government official websites or health and education institutions websites (88.7%), other health professionals (57.3%), and scientific articles (56.2%). The use of social media was reported by 54.1% of the students, while TV programs were information sources used by 39.9% of the students. The least used information sources were professors (38.1%) and family members (7.8%).

**Conclusions:** The respondents seem to be acquainted to seek information in reliable sources and also use social media as a gateway to keep updated about the pandemics. Therefore, knowing where the dental students seek information about COVID-19 might facilitate dental school directors to approach such public continuously, providing them with trustable information on different platforms.

## Introduction

COVID-19 is a virus-mediated disease caused by a member of the coronavirus family SARS-Cov-2 originated in Wuhan, Hubei, China in December 2019 [1,2]. A few months after the first cases, the disease has become pandemic, affecting more than 22 million people and causing over 782,000 deaths in 216 nations, according to the World Health Organization (WHO) COVID-19 dashboard on 21 August 2020 [3]. In Latin American countries, Brazil has risen to the spotlight of leading nations with the highest number of cases and deaths episodes due to COVID-19, ranking number 2 globally. Since the first diagnosed case in Brazil, there have been over 3,4 million cases confirmed along with staggering 111,100 numbers of death episodes by 21 August 2020 [3].

As the cases of COVID-19 increased globally and the disease pathophysiology has been constantly described, there has been an avalanche of information on this issue. By 21 August 2020, the keyword “COVID-19” yielded over 42,000 and 60,000 indexed articles on PubMed and Google Scholar, respectively, let alone over 6,5 billion results were acquired on Google search engine only in 2020. Even more impressive is the number of posts on social media platforms, which spread rapidly and easily [4,5]. This social media content has a high potential to carry on misleading information, hindering public health policies, and ultimately it can create a global epidemic of misinformation [6-9]. Recent reports revealed that individuals who get their news from social media are more likely to have misperceptions about COVID-19, whereas those who consume more traditional news media have fewer misperceptions and are more likely to follow public health recommendations like social distancing [10]. Along with this concerning findings, the information-seeking patterns can also modulate attitudes and behaviors towards this crisis [11].

Once the exposure to online health information has been associated with health-related behaviors in different populations and contexts [12-15], understanding the information-seeking behavior of dental students and their infosphere can be a necessary step toward building efficient educational planning in the context of COVID-19. These students are at direct contact with patients [16,17], and can funnel information to their niches. In this sense, educational institutions have such a role in fostering and providing the academic community with scientifically-oriented and official information to battle the current wave of infodemics so that dental students can be better prepared to tackle any related issue [16].

In Brazil, 350 tertiary institutions are responsible for formal dental education of up to 125,585 students across the country [18]. During the current university lockdown and social distancing, the e-learning regimen has taken place while hands-on experience has been discontinued for a while, profoundly affecting up to the totality of these students. However, considering social inequalities regarding information access (19,20), the overload of information regarding COVID-19 [9], and the infosphere as a behavioral modulator [15] it is necessary to identify the information source of dental students about COVID-19 as a tool to fight off misperceptions and misinformation among them, as well as to enforce educational policies toward this end. Thus, this study aimed to identify the source of information Brazilian dental students use regarding COVID-19.

## Methods

### Ethical aspects

This research protocol was approved by the research ethics committee of the School of Dentistry of Ribeirao Preto at the University of Sao Paulo (CAAE: 33608320.5.0000.5419). The study consists of a cross-sectional survey directed to a sample of dental students.

### Survey content

This study is part of a broader investigation and the details on how the questionnaire was developed and administered will be published elsewhere. In summary, a self-administered questionnaire about the awareness and knowledge of dental students about COVID-19 and its impact on the undergraduate dental practice was hosted online (Google Forms). The questionnaire contained 20 mandatory close-ended items, divided into four sections: 1) demographic and academic profile (n=6); 2) general knowledge about the COVID-19 (n=4); 3) knowledge about the preventive measures to avoid COVID-19 spread on the undergraduate dental practice (n=2); and 4 perception about the COVID-19 impacts on the undergraduate dental courses (8). The current work covers only the part of the 4^th^ section perceptions in which the students responded where or with whom they usually get information about COVID-19. Possible answers were government official websites or health and education institutions websites, TV Programs, professors, social media, scientific articles, other health professionals, and family members. The other data collected will be published elsewhere.

## Recruitment and data collection

According to data of the last Brazilian Tertiary Education Census [18], there are 125,585 undergraduate students enrolled in dentistry courses in Brazil, considering public and private institutions. All these students were eligible to participate in the research. Recruitment was conducted through an Instagram® (@covid.forp) (Facebook, Menlo Park, CA) social networking campaign, which started on July 4 and lasted until July 14. Details on how the recruitment was performed will be published elsewhere.

### Data analysis

The data collected were extracted from Google Forms and converted to Excel (Microsoft, USA) sheets. The frequencies distribution for the source of information about COVID-19 was analyzed by gender, age, and type of institution (public and private).

## Results

Table 1 depicts that the internet was the most common source of information of dental students about COVID-19, given that government official websites or health and education institutions websites were the answer more frequently chosen (88.7%). Interestingly, the students also referred to other health professionals as those with whom they seek information about the disease (57.3%). A very small proportion of dental students obtained their information from family members (7.8%). More than half of the students used scientific articles (56.2%) and social media (54.1%) as sources of information about COVID-19, a proportion that overcame the one for TV programs (39.9%) and professors (38.1%).

Both male (88.9%) and female (88%) sought information twice as much in official websites than in TV programs (40.3 and 38%, respectively) and with professors (37% and 42.2%, respectively) (Table 2). Moreover, male students used more scientific articles and less social media (59% and 50%, respectively) than female students (55.3% and 55.2%, respectively) (Table 3). Table 4 shows that official websites were more frequently used as sources of information by those who study dentistry in private institutions (91%), while the acquisition of information from scientific articles was more frequently used by students from public dental schools (60%). Moreover, comparable proportions of dental students from the public (54.2%) and private (54.1%) institutions used social media as an information source.

Table 4 displays that the use of official websites increased with age, being it absolute in those older than 39 years old. On the other hand, the use of social media decreased with age, dropping from 56.6% in the group of students younger than 25 years old to 20% in the group older than 39 years old. The use of scientific articles by students aging 25 to 32 years old was more frequent (68.6%) than by those aging between 18 to 25 years old (54.4%). Likewise, the use of scientific articles doubled in the group of dental students older than 39 years (87%) in comparison to the ones aging 25 to 32 years old (43.5%). TV programs were a source of information less frequently used by students older than 32 years old, and those older than 39 years old were less likely to obtain information with their family members.

Table 5 shows that the use of official websites increased as the year of undergraduate enrollment increased, reaching 91.9% in the group of students from the 5^th^ year of dental school. On the other hand, the frequency of students who had professors as information sources decreased as the time of enrollment increased from the third to the fifth year of dental school. Moreover, while 52.1% of last-year dental students used social media to stay informed about COVID-19, only 30.8% of them saw their professors as someone with whom they can obtain information.

## Discussion

The outbreak of COVID-19 has challenged individuals, communities, and educational institutions given the need for social distancing and due to the sanitary measures imposed by the pandemics [2,21]. Health care students and professionals have been strongly affected given the imminent risk of infection spread associated with their process of learning and working [22-25]. As dentists are at the top of professionals at risk [22,26], so are dental students, who have been facing a hard time trying to complete their education during these uncertain times [16,27-30]. As it has been demonstrated, the course of infection control can be shaped by how governments enact timely policies and disseminate information [11]. Thus, here we investigated where Brazilian dental students usually seek information about COVID-19. These data might aid dental schools in choosing the best platforms to display educative campaigns.

Firstly, we observed that regardless of gender, age, type of institution, and year of enrollment, dental students had government official websites or health and education institutions websites as their main source of information about COVID-19. As far as we know, this is the first study to investigate information seeking about COVID-19 by dental students. As dental schools in Brazil offer dental undergraduate courses to over 125,000 students [18], knowing that they are familiarized with getting informed on official websites might reveal the students’ precaution in getting information through reliable online platforms. In this context, data of the national portals of the 193 United Nations Member States showed that by 8 April 2020, around 86 percent of nations (167 countries) had included information and guidance about COVID-19 in their portals [31]. However, it has been shown that a more advanced strategy is having a dedicated portal or section about the COVID-19 [11]. Thus, we suggest that dental schools could display a COVID-19 page on their school’s website, where they could provide information about preventive measures and on the statistics about the outbreak, focusing on the local situation of the city and campus [17]. Such information helps people make informed decisions about their daily routines and build public trust [11].

Contrary to what we expected, social media did not figure among the top source of information used by dental students. Such results are also contrary to what other investigations with university students have been showing [32]. As the use of social media in Brazil increased significantly during the social distancing period [33], also because we recruited the respondents via Instagram, we expected that such tools would be more frequently reported by the students as information sources. Even not being at the top of the most used information source, more than half of the respondents used social media to get informed about the pandemics, mainly the younger ones. These results are an alert to dental school directors to the importance of social media in disseminating information to the students, especially in cases of pandemics. In this scenario, it is concerning the fast and uncontrolled spread of news, which might be associated with misinformation [6,7,9,11,13,15,34]. Moreover, social media users are more likely to believe false information. For instance [10], manipulation of information with doubtful intent might be amplified through social networks, spreading farther and faster like a virus, the so-called infodemics [11,34]. Thus, an effort should be made by dental schools to keep their social media activity and updated to provide their students with trustable information, fighting fake news [17].

Interestingly, we also observed that the respondents considered other health professionals as someone with whom they obtain information. Such data represents the acknowledgment of the critical role of healthcare workers during pandemics, as it has also been stressed by health care authorities [35]. Surprisingly, professors were one of the least chosen sources of information of the dental students, which might be associated with the social distancing imposed by the pandemics. In this aspect, it is known that COVID-19 has strongly impacted teaching and learning in dental schools, which had to move online, challenging the interaction among students and professors [17,36,37]. Corroborating with which has been shown to other university students, family members at the bottom of the list when evaluating with whom the students get information about COVID-19 [32]. Moreover, as it has been shown elsewhere, university students seem to seek information in scientific publications [38], as we observed in our sample of dental students. In this regard, according to the Nature Index, there have been published 67,753 scientific publications about COVID-19 [39]. As the rise in publications represents the massive effort of scientists to overcome the pandemic crisis, it also signifies that it is becoming difficult to follow all daily updates, mainly for undergraduate students, who are at the beginning of their academic life. Thus, we reinforce the role dental schools in funneling the available information, providing the students with reliable sources and leading them to a safer return to hands-on activities.

## Conclusions and final considerations

Contrary to other studies with university students, the most used information sources of the Brazilian dental students who participated in our research where official government and educational websites, followed by other health professionals and scientific articles. However, the use of social media was also reported by a high proportion of the respondents. As a final remark, we emphasize that knowing where the dental students seek information about COVID-19 might facilitate dental school directors to approach such public continuously, providing them with trustable information on different platforms. Once it has been shown that when individuals face risks, they seek information to reduce uncertainty, dental schools in countries such as Brazil, where the epidemic is rocketing in cases and deaths, should implement strategies to keep their students updated. Such precaution would provide students with knowledge, guiding them to proper attitudes.

## Data Availability

All data are available upon request to the corresponding author

## Conflics of interest

The authors declare no conflics of interest

